## Supplementary file for "Rural prioritization may increase the impact of COVID-19 vaccines in Sub-Saharan Africa due to ongoing internal migration: A modeling study"

### 1 Age pyramid by urban and rural

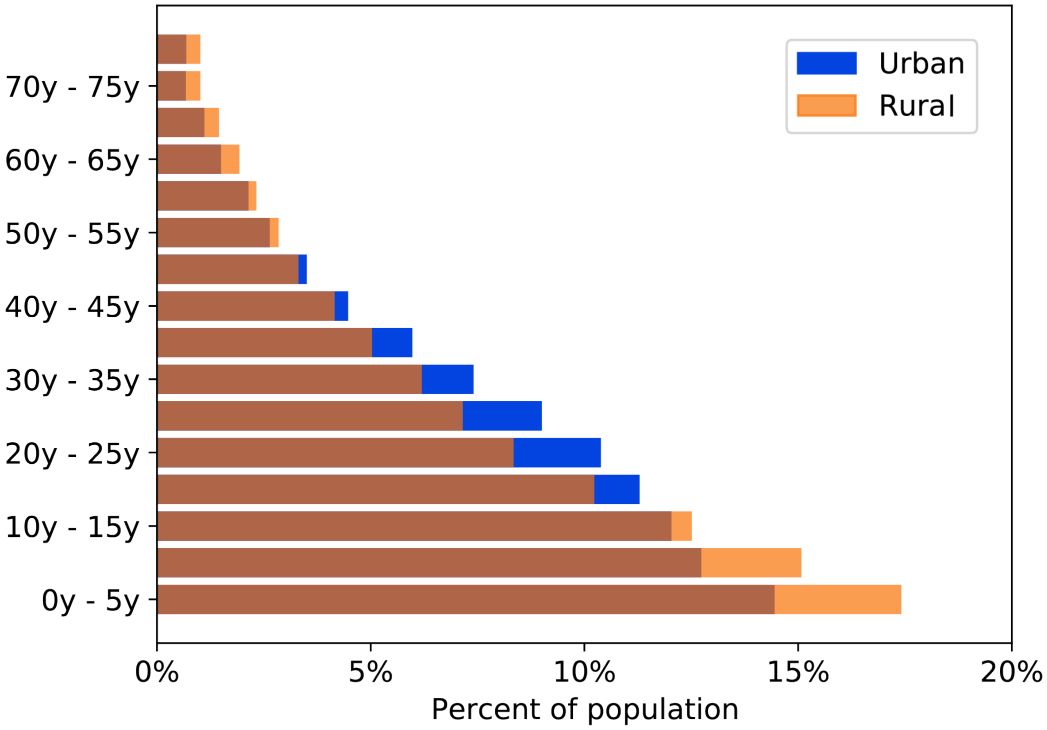


Figure 1: Age pyramid by urban and rural averaged over 37 sub-Saharan countries.

#### 2 Contact matrices by urban and rural

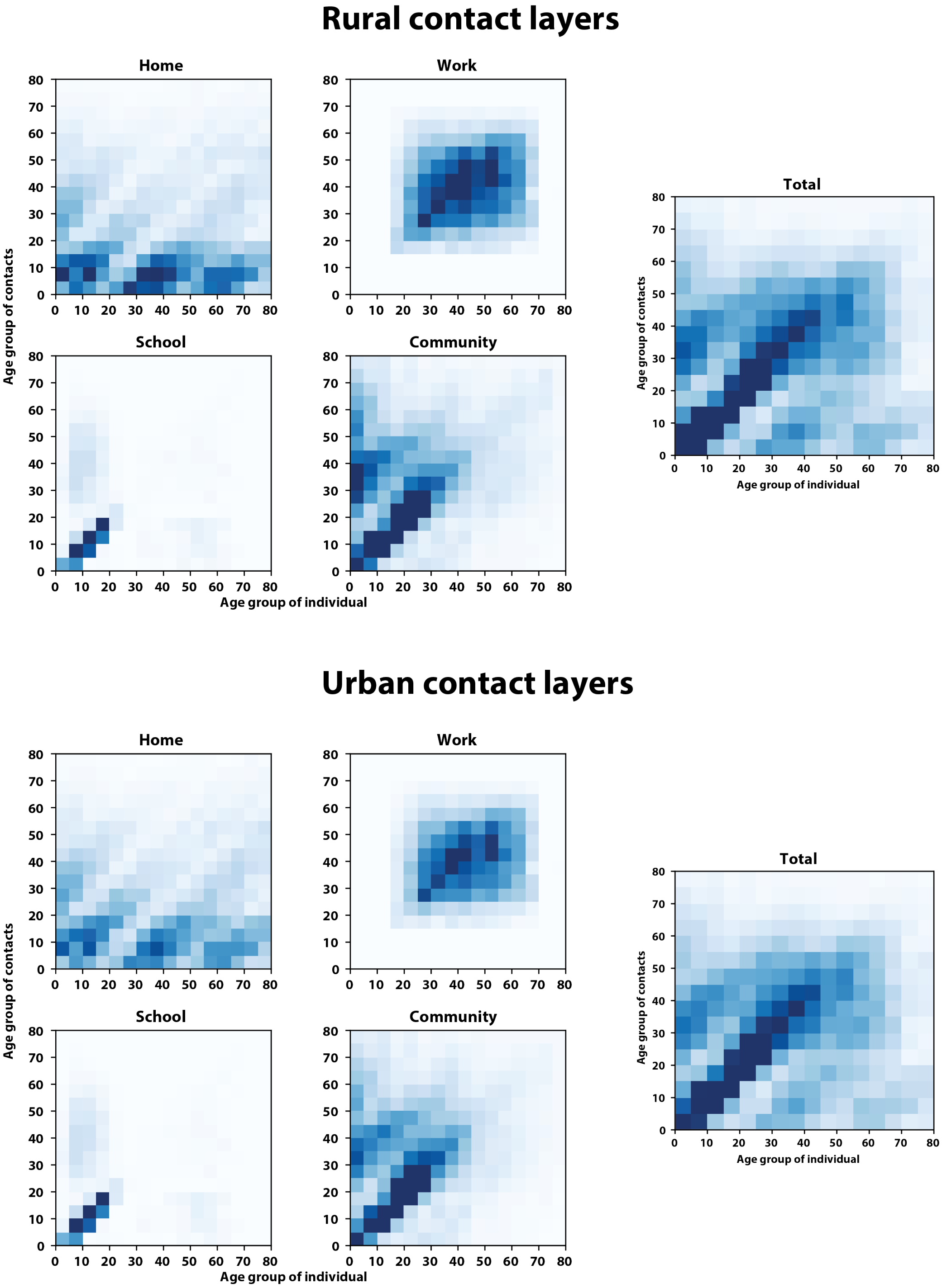


Figure 2: Contact layers by urban and rural averaged over 37 sub-Saharan countries. The total contact layer is obtained by summing contacts from home, work, school and community settings.

### 3 Prioritization by age

#### 3.1 Low migration and R0 = 2.0

Vaccination campaign impacts by age prioritization. Vaccination could begin with young adults (15 to 50 years old), old adults (70 years old and older), or without priority age groups (random).

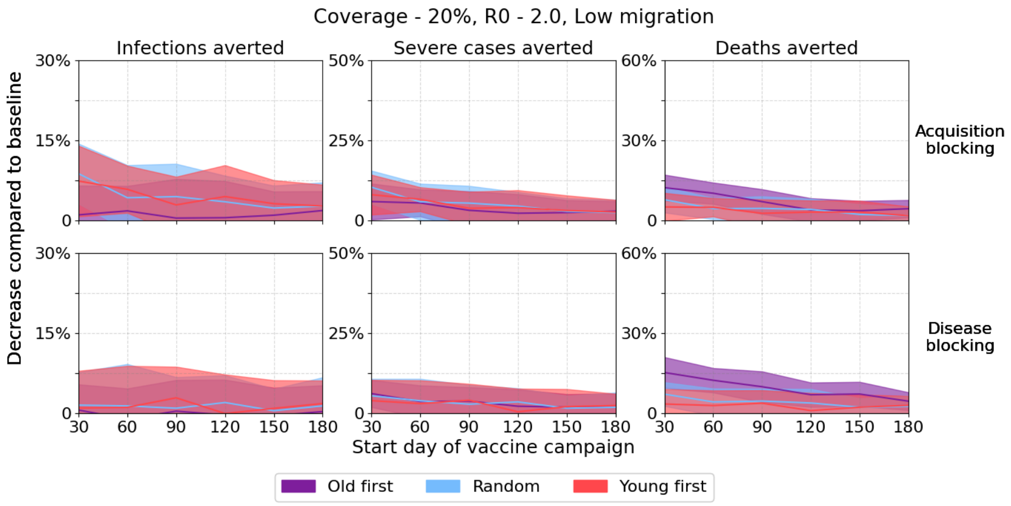


Figure 3: Low migration, R0=2.0, 20% coverage.

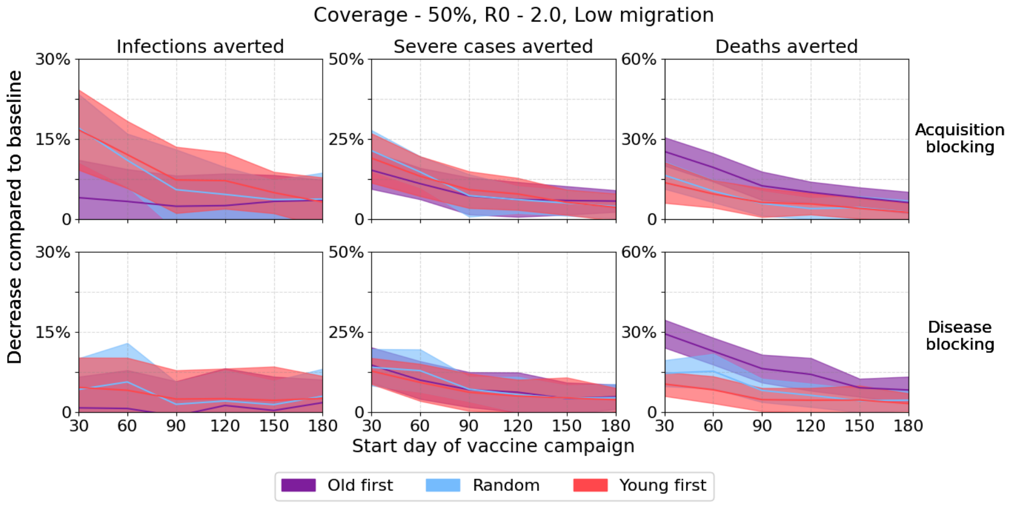


Figure 4: Low migration, R0=2.0, 50% coverage.

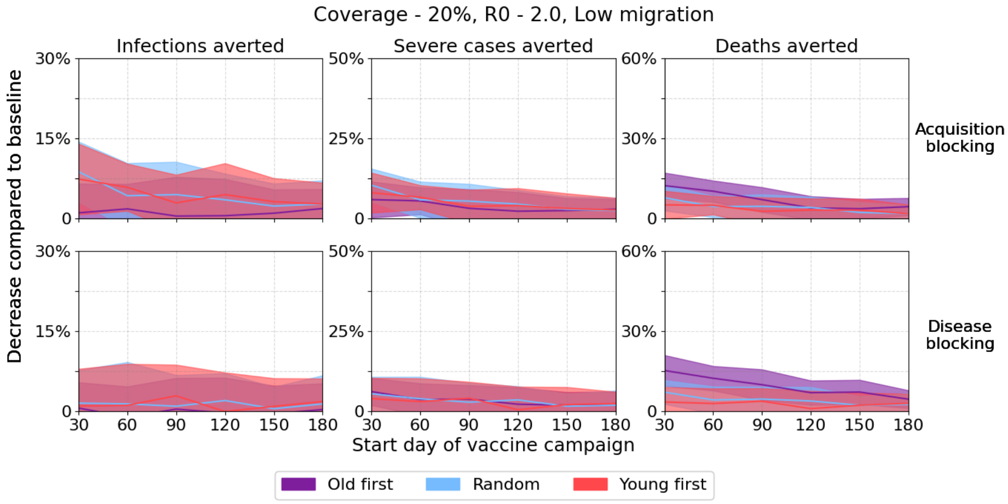


Figure 5: Low migration, R0=2.0, 80% coverage.

#### 3.2 Low migration and R0 = 2.4

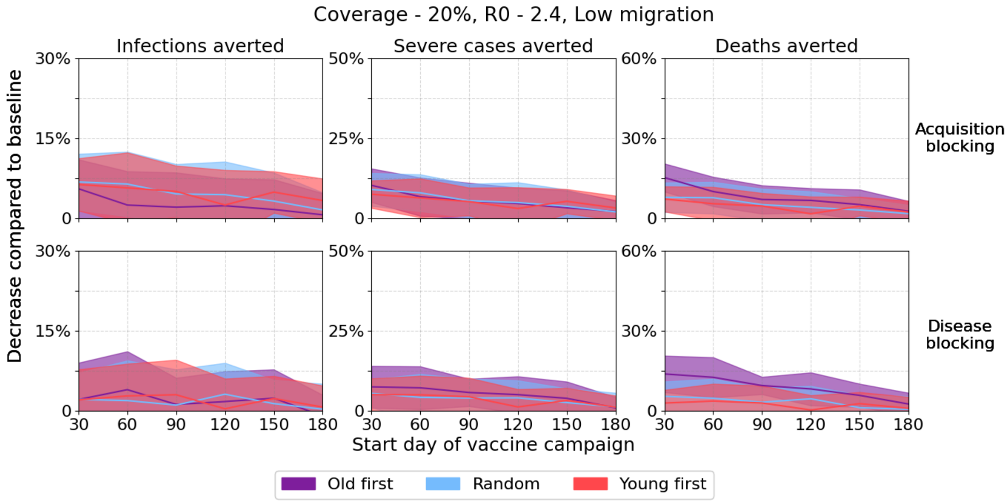


Figure 6: Low migration, R0=2.4, 20% coverage.

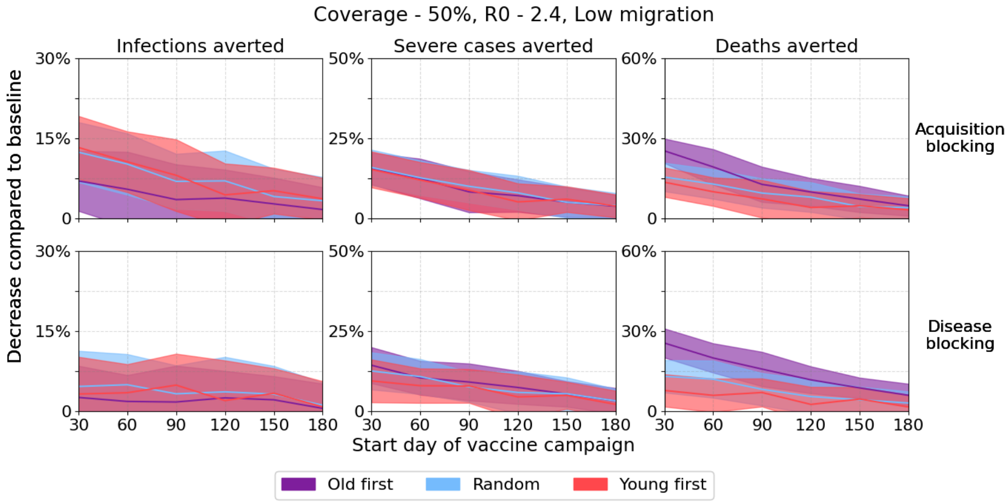


Figure 7: Low migration, R0=2.4, 50% coverage.

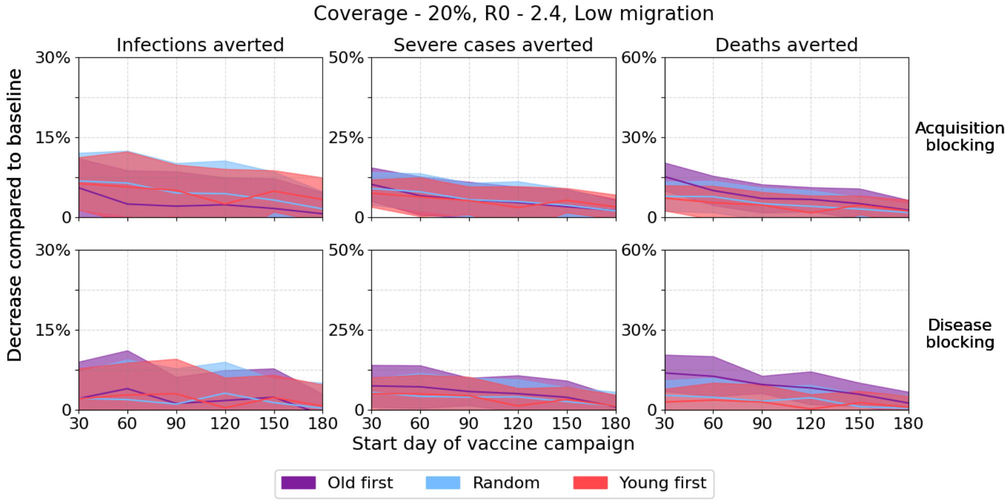


Figure 8: Low migration, R0=2.4, 80% coverage.

#### 3.3 Low migration and R0 = 2.8

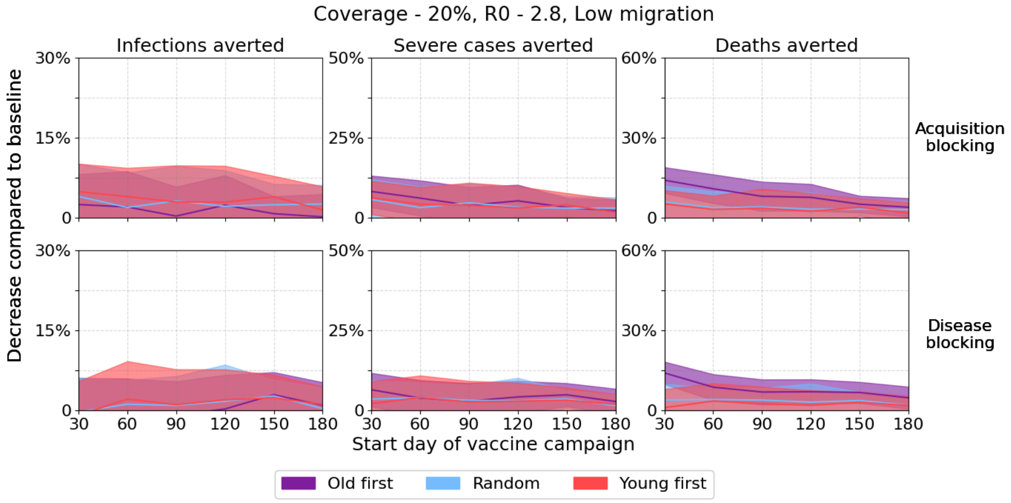


Figure 9: Low migration, R0=2.8, 20% coverage.

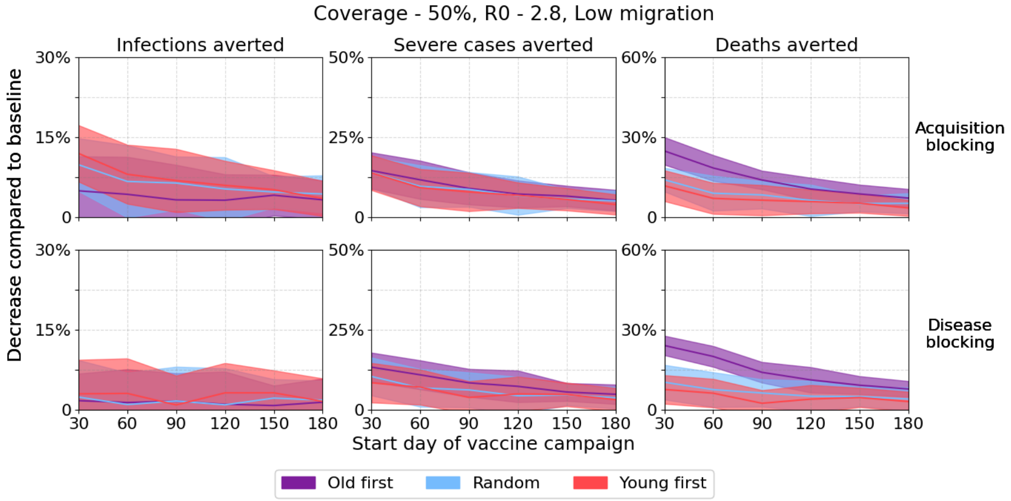


Figure 10: Low migration, R0=2.8, 50% coverage.

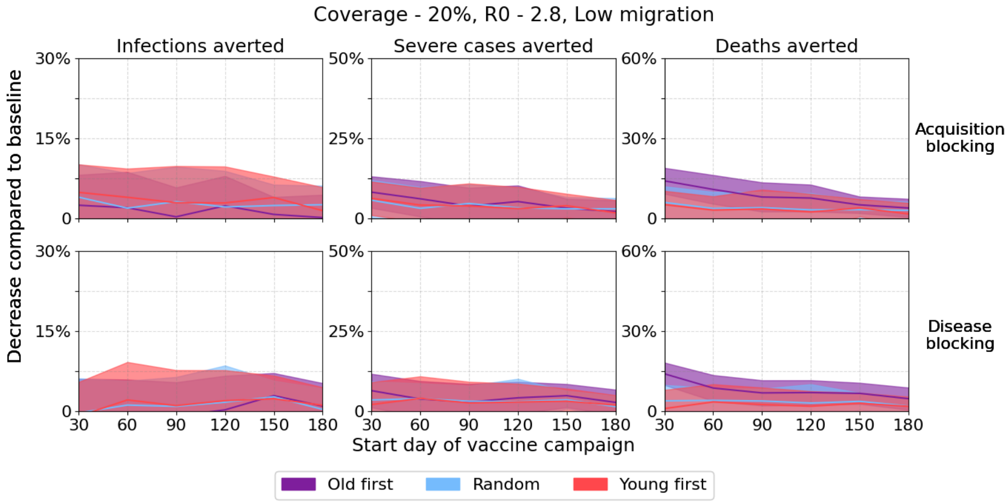


Figure 11: Low migration, R0=2.8, 80% coverage.

#### 3.4 Medium migration and R0 = 2.0

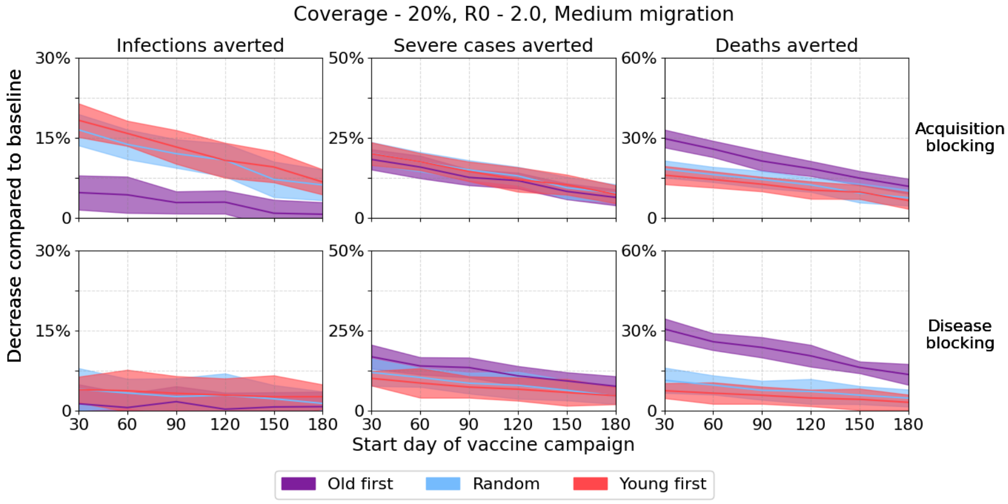


Figure 12: Medium migration, R0=2.0, 20% coverage.

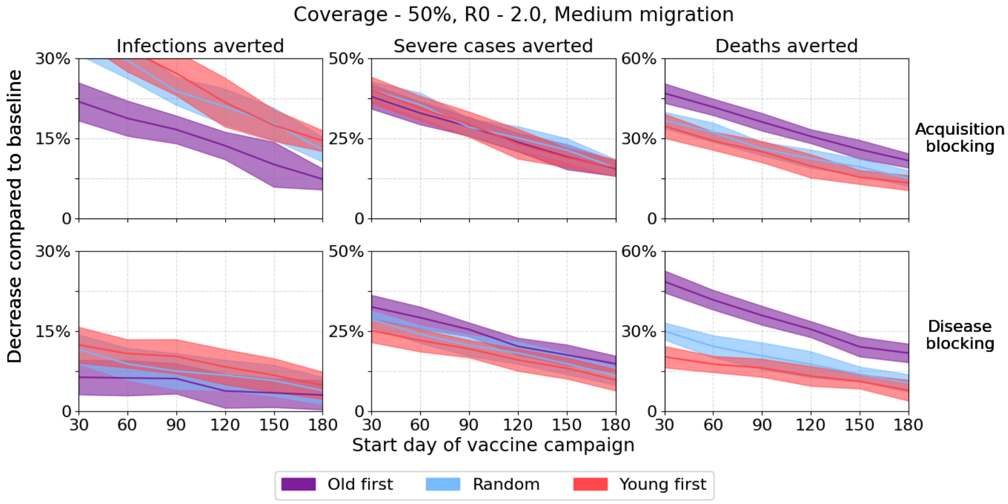


Figure 13: Medium migration, R0=2.0, 50% coverage.

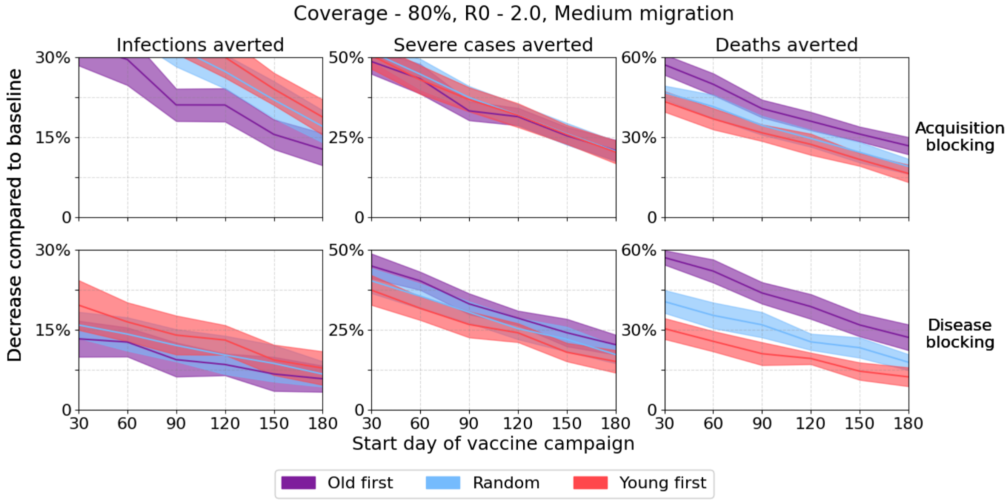


Figure 14: Medium migration, R0=2.0, 80% coverage.

#### 3.5 Medium migration and R0 = 2.4

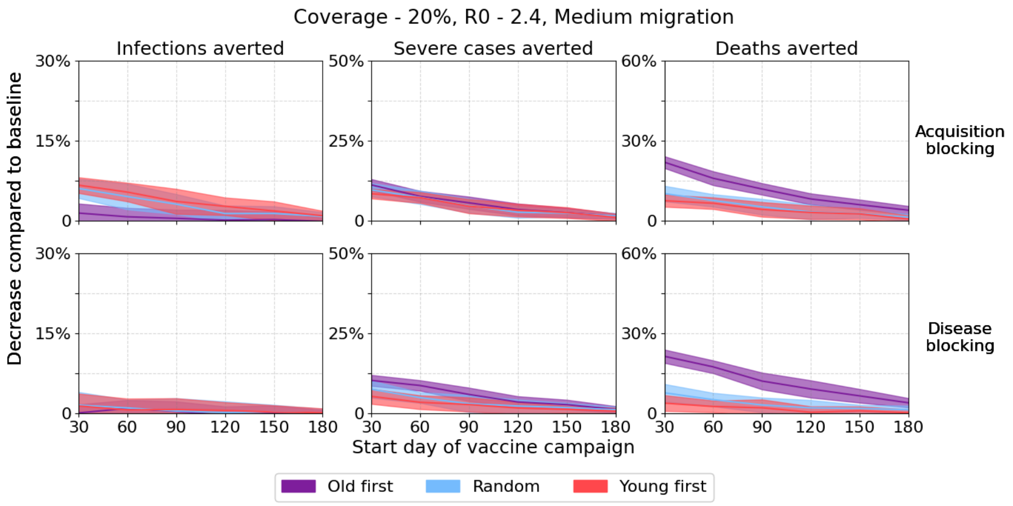


Figure 15: Medium migration, R0=2.4, 20% coverage.

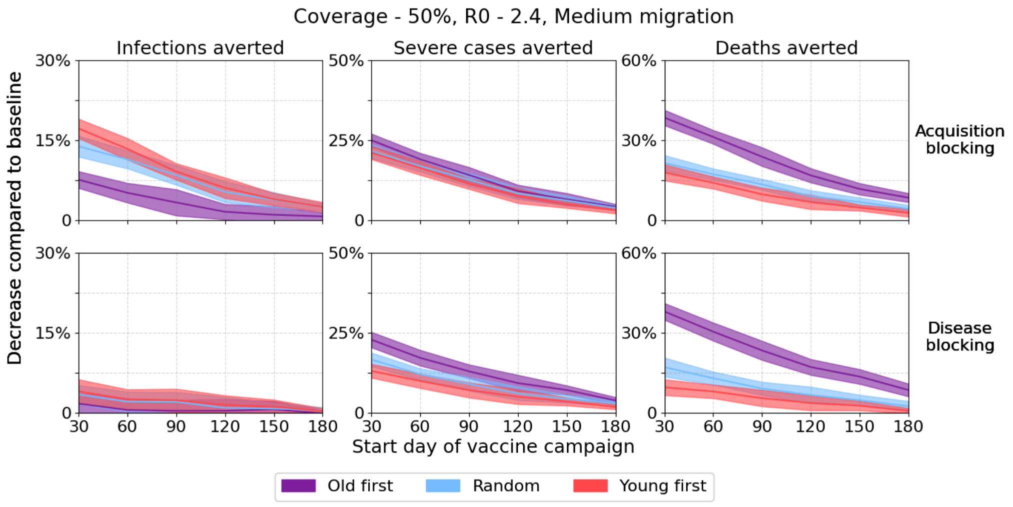


Figure 16: Medium migration, R0=2.4, 50% coverage.

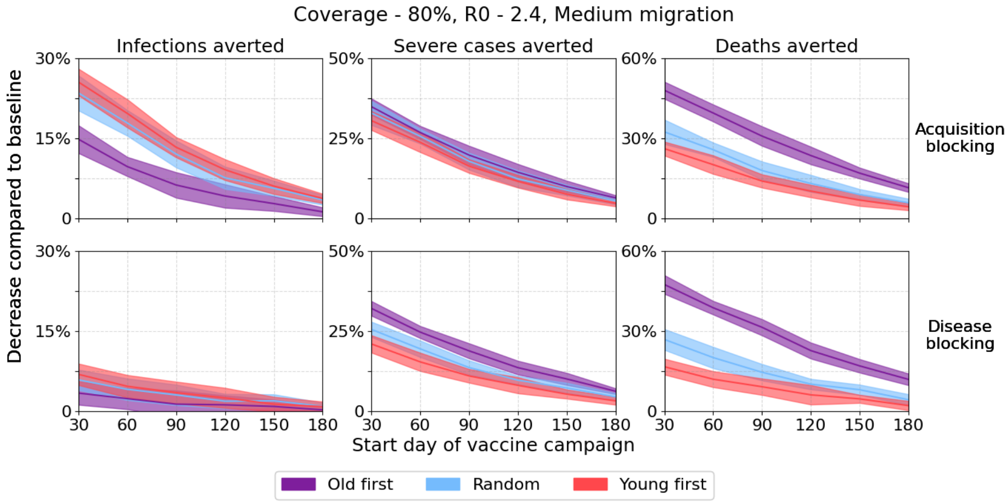


Figure 17: Medium migration, R0=2.4, 80% coverage.

#### 3.6 Medium migration and R0 = 2.8

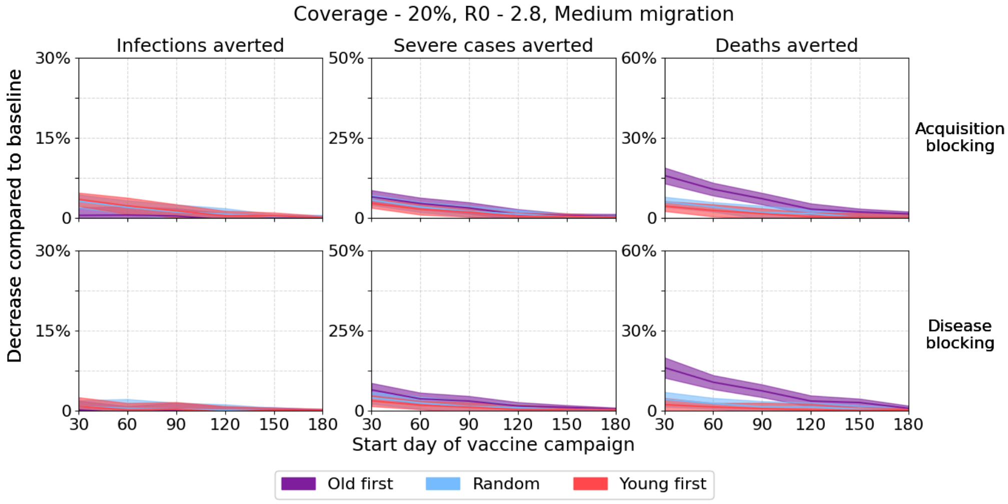


Figure 18: Medium migration, R0=2.8, 20% coverage.

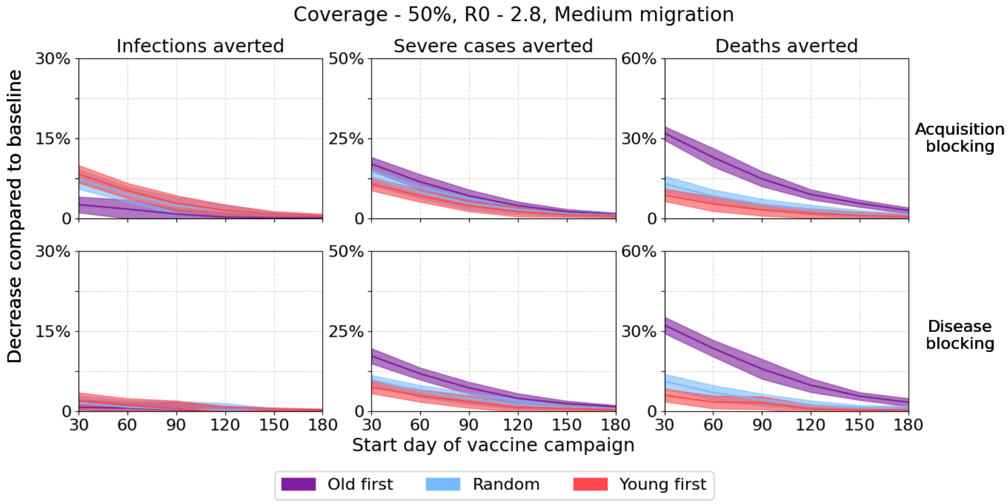


Figure 19: Medium migration, R0=2.8, 50% coverage.

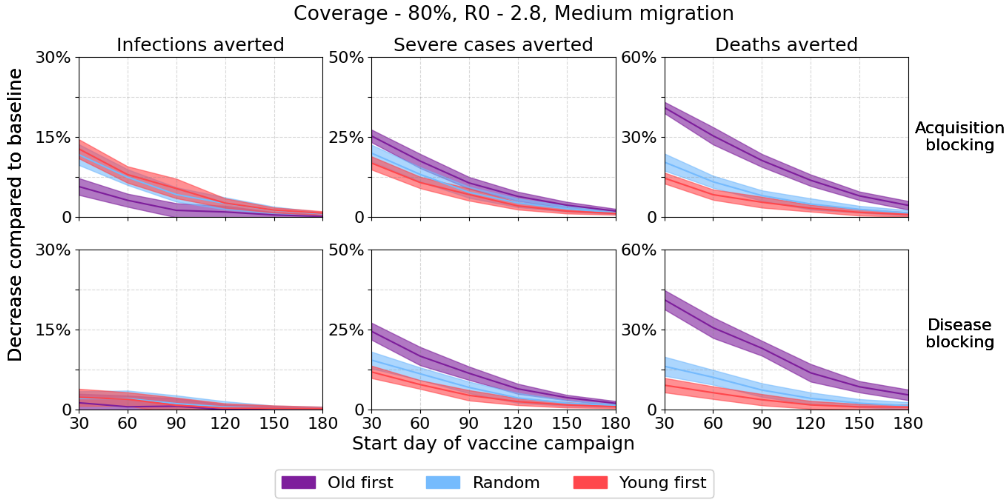


Figure 20: Medium migration, R0=2.8, 80% coverage.

#### 3.7 High migration and R0 = 2.0

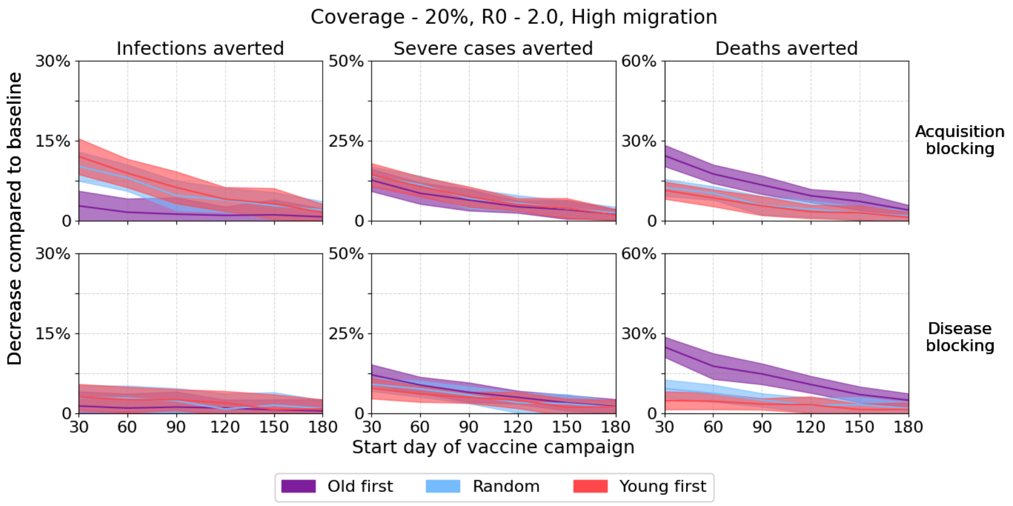


Figure 21: High migration, R0=2.0, 20% coverage.

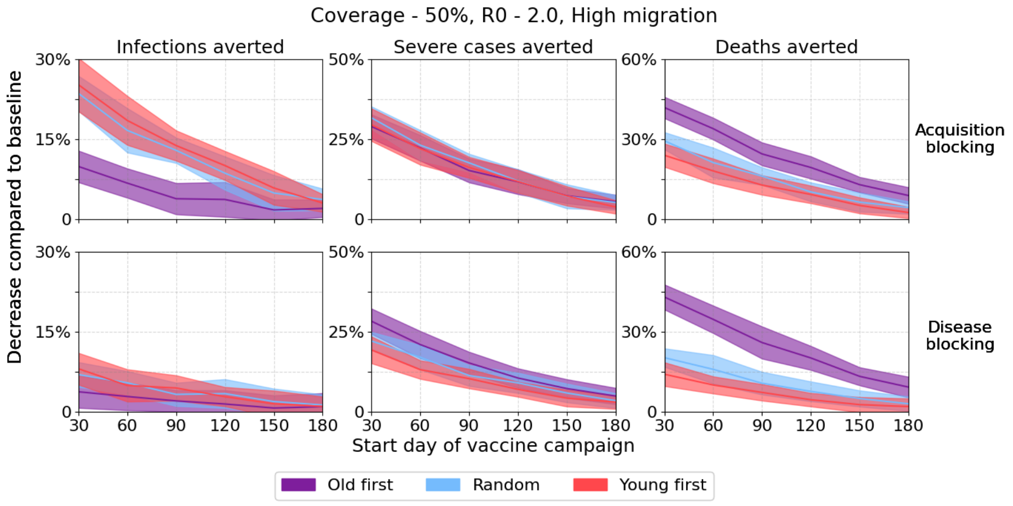


Figure 22: High migration, R0=2.0, 50% coverage.

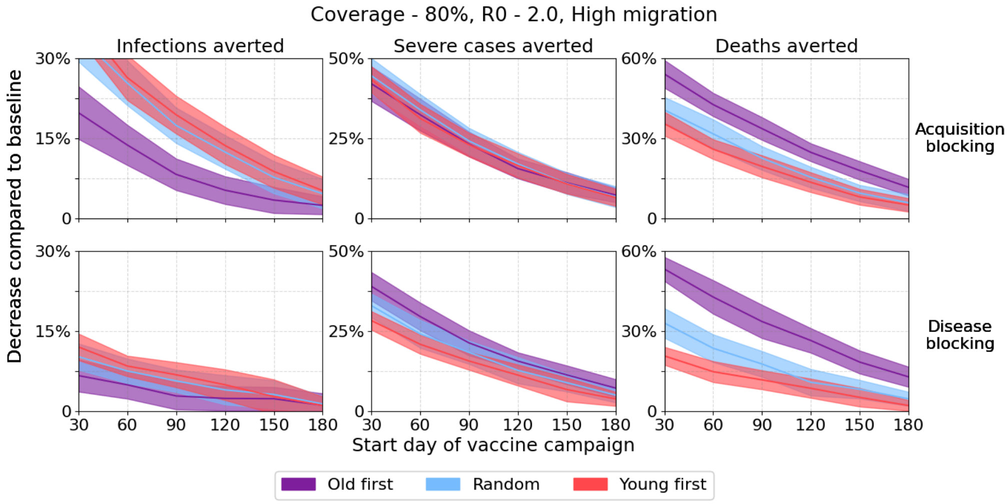


Figure 23: High migration, R0=2.0, 80% coverage.

#### 3.8 High migration and R0 = 2.4

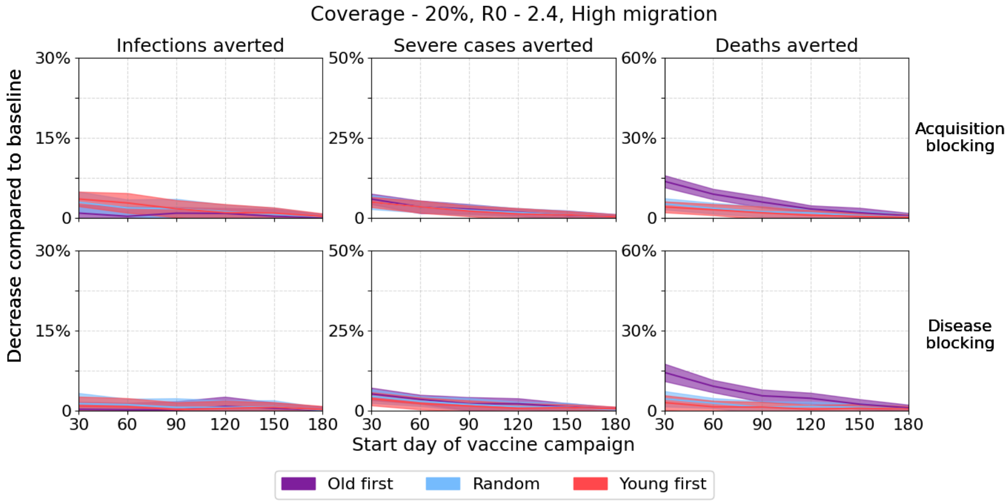


Figure 24: High migration, R0=2.4, 20% coverage.

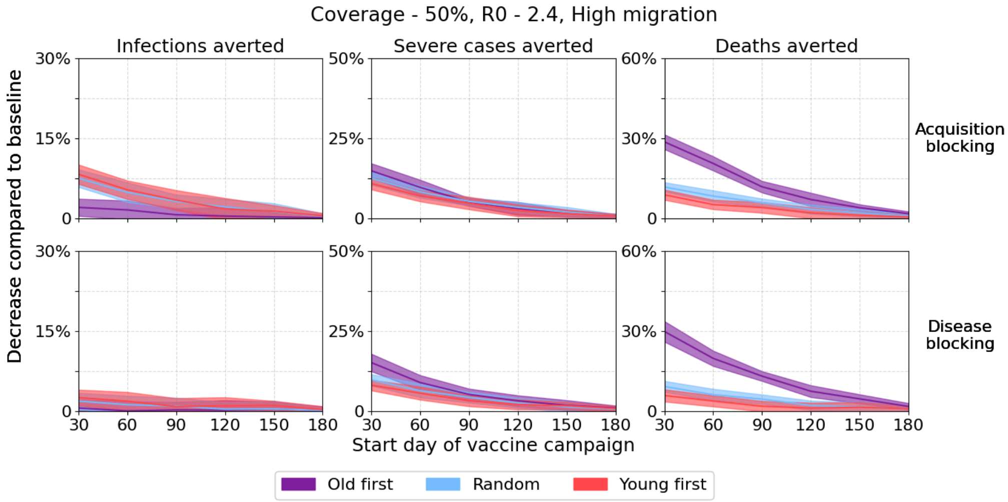


Figure 25: High migration, R0=2.4, 50% coverage.

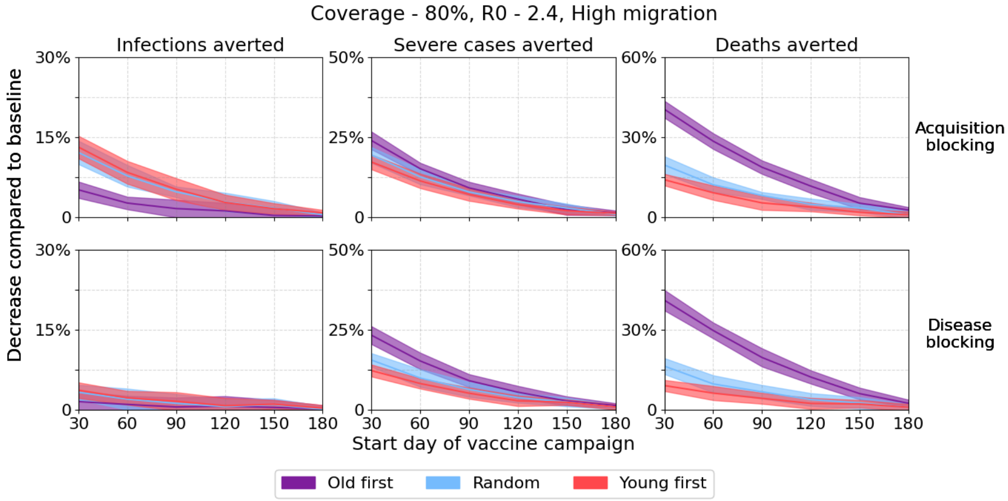


Figure 26: High migration, R0=2.4, 80% coverage.

#### 3.9 High migration and R0 = 2.8

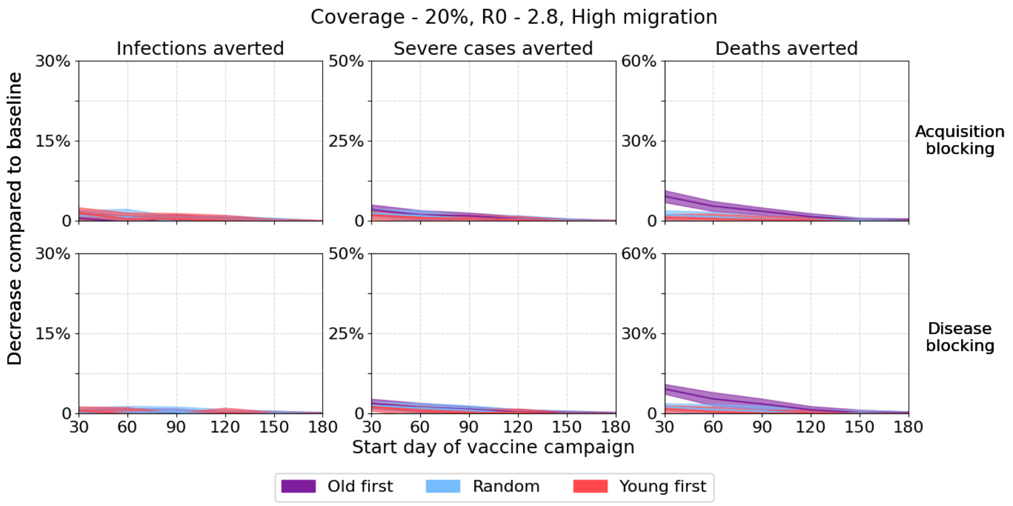


Figure 27: High migration, R0=2.8, 20% coverage.

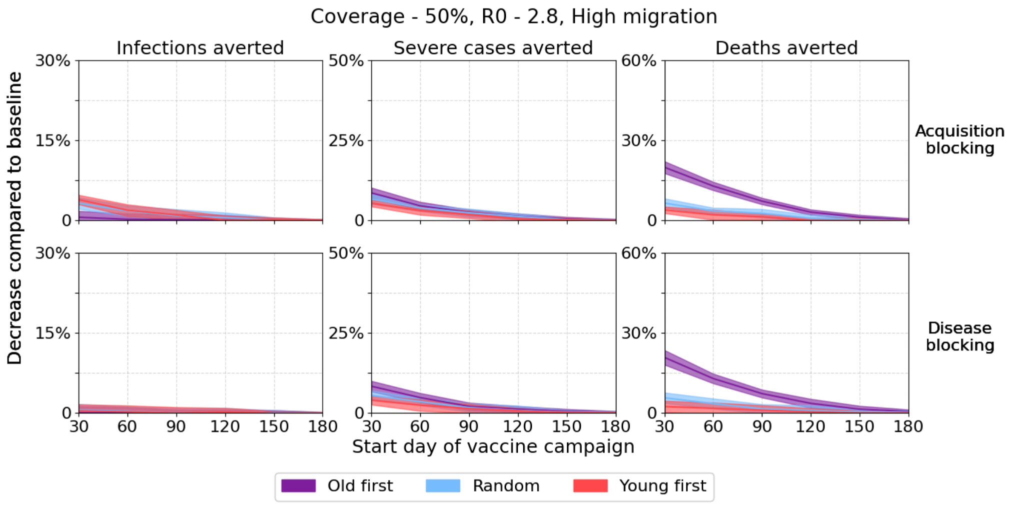


Figure 28: High migration, R0=2.8, 50% coverage.

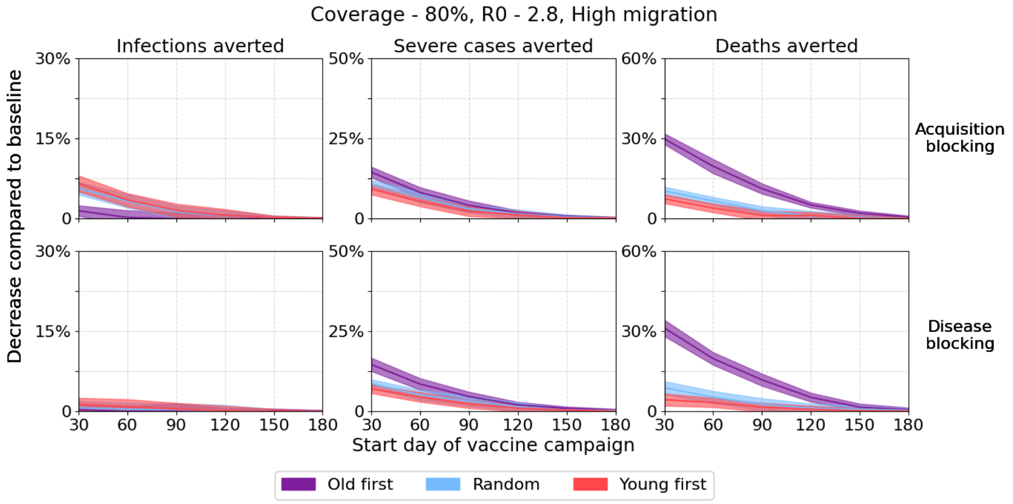


Figure 29: High migration, R0=2.8, 80% coverage.

### 4 Spatial prioritization

#### 4.1 Acquisition blocking

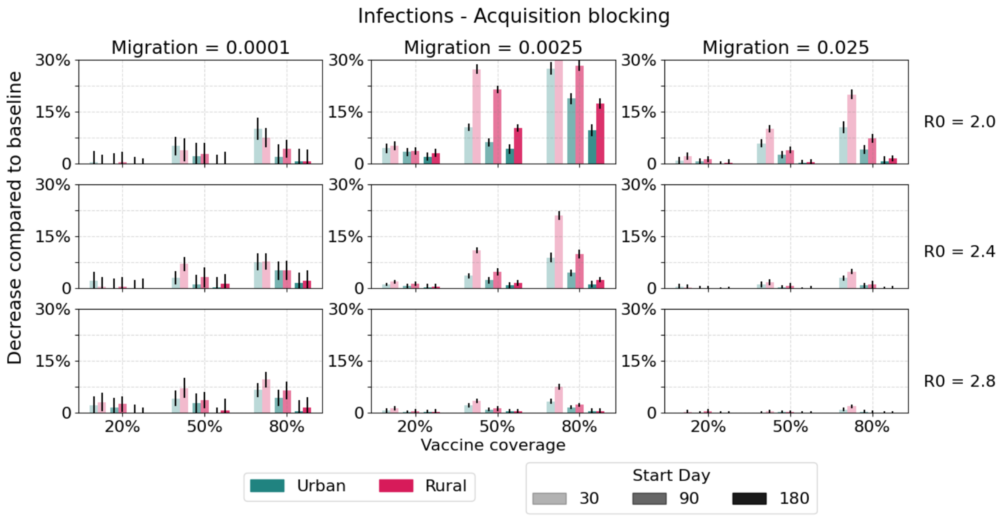


Figure 30: Infections averted via spatial prioritization using an acquisition blocking vaccine.

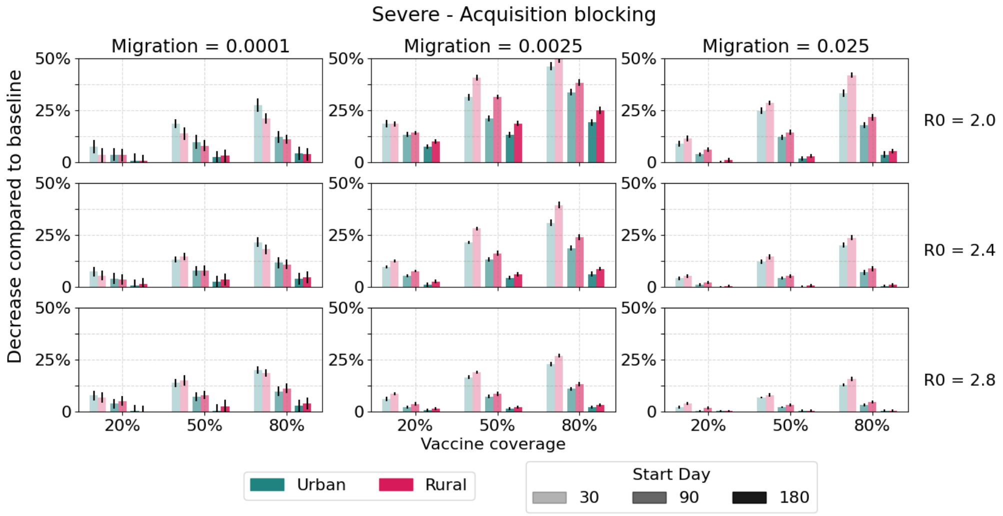


Figure 31: Severe cases averted via spatial prioritization using an acquisition blocking vaccine.

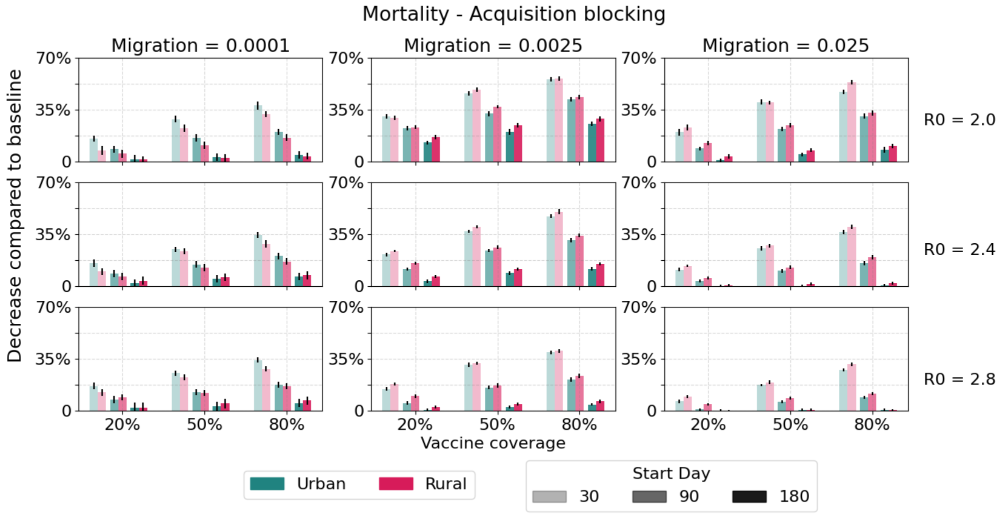


Figure 32: Deaths averted via spatial prioritization using an acquisition blocking vaccine.

#### 4.2 Disease blocking

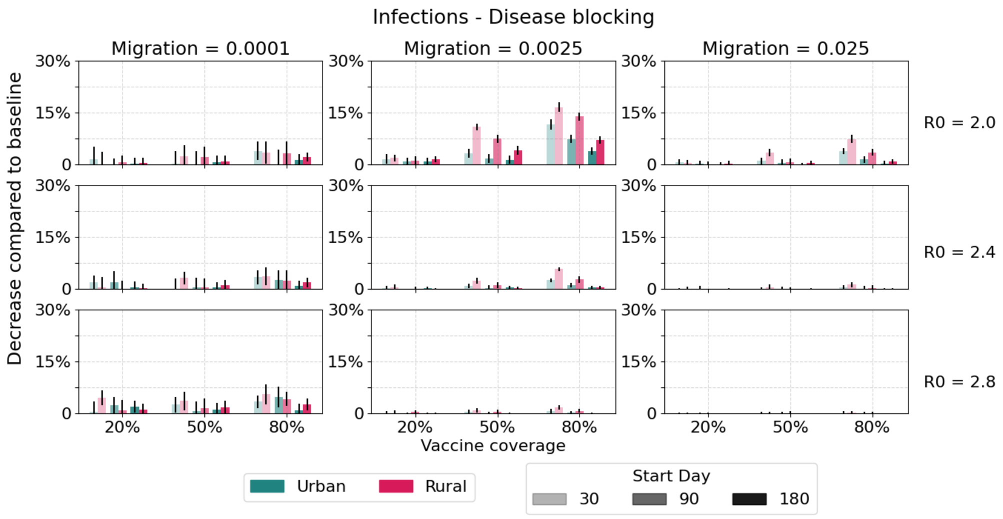


Figure 33: Infections averted via spatial prioritization using an disease blocking vaccine.

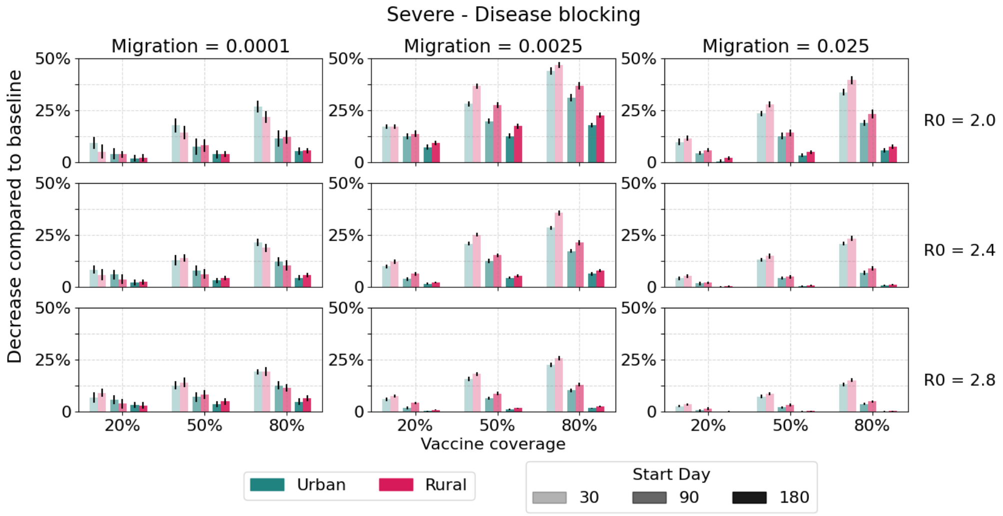


Figure 34: Severe cases averted via spatial prioritization using an disease blocking vaccine.

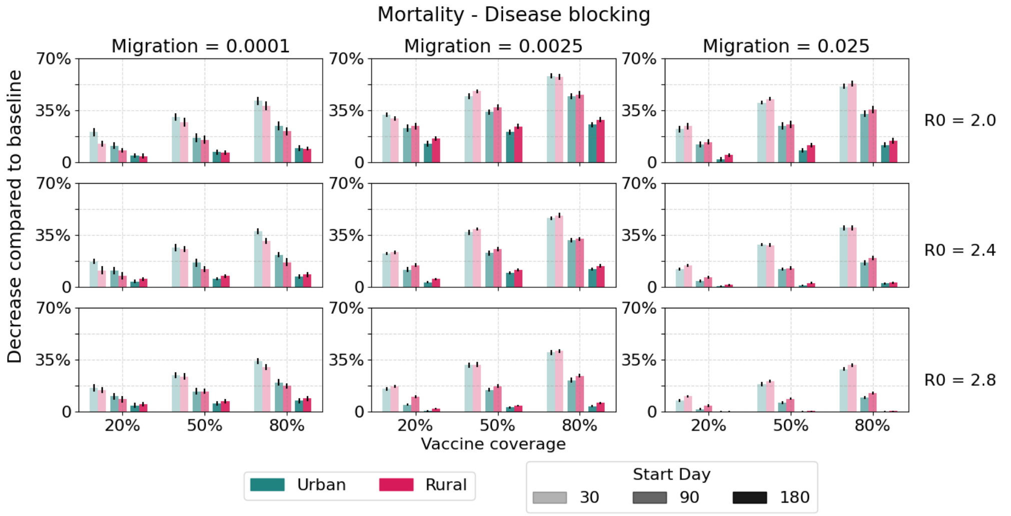


Figure 35: Deaths averted via spatial prioritization using an disease blocking vaccine.
